# Smartphone Passive Digital Phenotyping in Frontotemporal Lobar Degeneration

**DOI:** 10.64898/2026.09.18.26363439

**Authors:** Emily W Paolillo, Sreya Dhanam, Jack Carson Taylor, Mark Sanderson-Cimino, Ray Fregly, Rowan Saloner, Kaitlin B Casaletto, Joel H Kramer, Bruce L Miller, William W Seeley, Maria Luisa Gorno-Tempini, Peter A Ljubenkov, Julio C Rojas, Suzee Lee, Virginia Sturm, Brian Appleby, Ece Bayram, David Clark, Ciaran M Considine, Richard R Darby, Gregory S Day, Alyssa De Vito, Mark Eldaief, Julie A Fields, Nupur Ghoshal, Edward D Huey, David J Irwin, Kejal Kantarci, Justin Y Kwan, Ian R Mackenzie, Joseph C Masdeu, Chiadi U Onyike, Alexander Pantelyat, Belen Pascual, Tanav Popli, Katya Rascovsky, Neguine Rezaii, Sonja W Scholz, Allison Snyder, M. Carmela Tartaglia, Bryan J Traynor, Bonnie Wong, John Kornak, Walter K Kremers, Leah K Forsberg, Hilary W Heuer, Bradley F Boeve, Howard J Rosen, Adam L Boxer, Adam M Staffaroni, the ALLFTD Consortium

## Abstract

**Introduction:** Frontotemporal lobar degeneration (FTLD) is a devastating disease that commonly results in early onset dementia, yet its rarity and heterogeneity limit large-scale research and clinical trials. Remote monitoring via low-burden digital health tools may overcome these barriers. We evaluated clinical utility of passive smartphone monitoring in FTLD using a multi-domain mobile assessment platform.

**Methods:** Participants were 567 adults (53% clinically normal, 21% prodromal FTLD, 26% symptomatic FTLD) who completed an in-person study visit and downloaded the ALLFTD Mobile App on their personal smartphones. The app delivered unsupervised cognitive tests and passively collected continuous data on battery percentage (proxy for smartphone use) and step count. Longitudinal follow-up included smartphone monitoring and annual in-person study visits. Primary analyses examined associations between passive smartphone features and markers of disease severity at baseline and longitudinally, and tested whether passive features added incremental value beyond app-based cognitive testing.

**Results:** Passive smartphone features were feasible to collect and showed excellent reliability with <2 weeks of monitoring (ICCs>0.9). Features capturing smartphone use and movement demonstrated sensitivity to gold-standard clinical measures of disease severity both at baseline and longitudinally. A classification model combining passive features, the app-based cognitive composite score, and demographics detected longitudinal functional decline (AUC=0.89); passive features alone (AUC=0.84) performed comparably to a cognitive screener (AUC=0.85).

**Discussion:** Passive smartphone monitoring is a valid, zero-burden measure of disease severity and progression in FTLD that adds modest incremental value beyond app-based cognitive testing. Findings support its use as a scalable digital endpoint in longitudinal FTLD research.

## BACKGROUND

As the global population ages, the growing prevalence and burden of neurodegenerative disease is increasingly characterized as a public health crisis (1–3). While there have been many recent advancements in disease modifying therapies for patients with Alzheimer’s disease (AD) (4), substantial barriers remain in frontotemporal lobar degeneration (FTLD) research that slow progress on clinical trials and translation of early research results into clinical use (5–8).

Although FTLD is a common cause of early-onset dementia, it is much rarer than AD overall (9), making recruitment and enrollment of participants with FTLD into research challenging, especially across ethnoculturally diverse populations (10). Further, FTLD is pathologically and clinically heterogeneous, as it is an umbrella term for a group of distinct proteinopathies that cause rare dementia syndromes typically characterized by impairments in behavior, language, and/or motor function (11). This clinical heterogeneity complicates our ability to measure and monitor multi-domain symptomatic changes that occur as the disease progresses; this reality challenges efforts to develop decentralized clinical trials that leverage remote assessments to overcome enrollment barriers, increase access, and reduce participant burden (12).

Digital health tools are increasingly being utilized for remote cognitive and functional assessment in aging patients with and without neurodegenerative disease (13, 14). Several smartphone-based cognitive assessment tools have been validated in individuals with AD (15), but these typically focus on domains primarily affected in AD, such as memory (16–19). To date, only one smartphone-based assessment platform has been validated in individuals with FTLD (20, 21). This ALLFTD Mobile app, developed in collaboration with Datacubed Health, delivers multi-domain FTLD assessments on participants’ own personal smartphones and has been deployed in the large multi-site U.S.-based ARTFL–LEFFTDS Longitudinal Frontotemporal Lobar Degeneration (ALLFTD) study (https://www.allftd.org) (20, 22, 23). The app was found to be feasible and acceptable in FTLD (24), and includes reliable, valid, and sensitive measures of executive functioning and memory (20, 25). Validation of its speech/language, motor, and gait tests in FTLD is currently underway (26, 27).

Because the ALLFTD Mobile App is a native app downloaded onto participants’ smartphones, it can also passively collect sensor data characterizing overall phone use and movement (20, 28). These data support digital phenotyping, defined as “the moment-by-moment quantification of the individual-level human phenotype *in situ* using data from personal digital devices” (29). Given the near-universal adoption of smartphones worldwide (30), digital phenotyping is increasingly recognized as a powerful method for comprehensive, unobtrusive, and individualized health monitoring, as it can capture continuous naturalistic behaviors in real-world environments with no added patient burden (31). In a disease where behavioral and functional changes are difficult to capture during brief clinic visits, passively collected smartphone data are a promising complement to the multi-domain active mobile assessments being validated for use in FTLD (32).

In this study of over 500 participants with and without FTLD, we examined whether passive smartphone monitoring can measure and track disease severity. We focused on features characterizing average and day-to-day variability in smartphone use, captured by proxy estimate of total smartphone battery drainage per day (28), and step counts, sourced directly from each phone’s built-in step tracker (**Fig. 1**). Our study primarily aimed to address validity and clinical utility in FTLD by: 1) determining associations between passive smartphone features and established clinical measures of cognitive and functional impairment at baseline (i.e., the first 30 days of passive smartphone monitoring); 2) testing whether baseline passive smartphone features prognosticate longitudinal clinical change; 3) examining whether changes in passive smartphone features track with concurrent change in established clinical measures over annual follow-up visits; and 4) evaluating the incremental value of passive smartphone features in addition to smartphone cognitive testing for longitudinal monitoring. As a prerequisite, we also first established feasibility of passive smartphone data collection, evaluated its reliability and the minimum monitoring length needed for reliable estimates, and determined agreement between smartphone use features and self-reported use.

**Figure 1.**
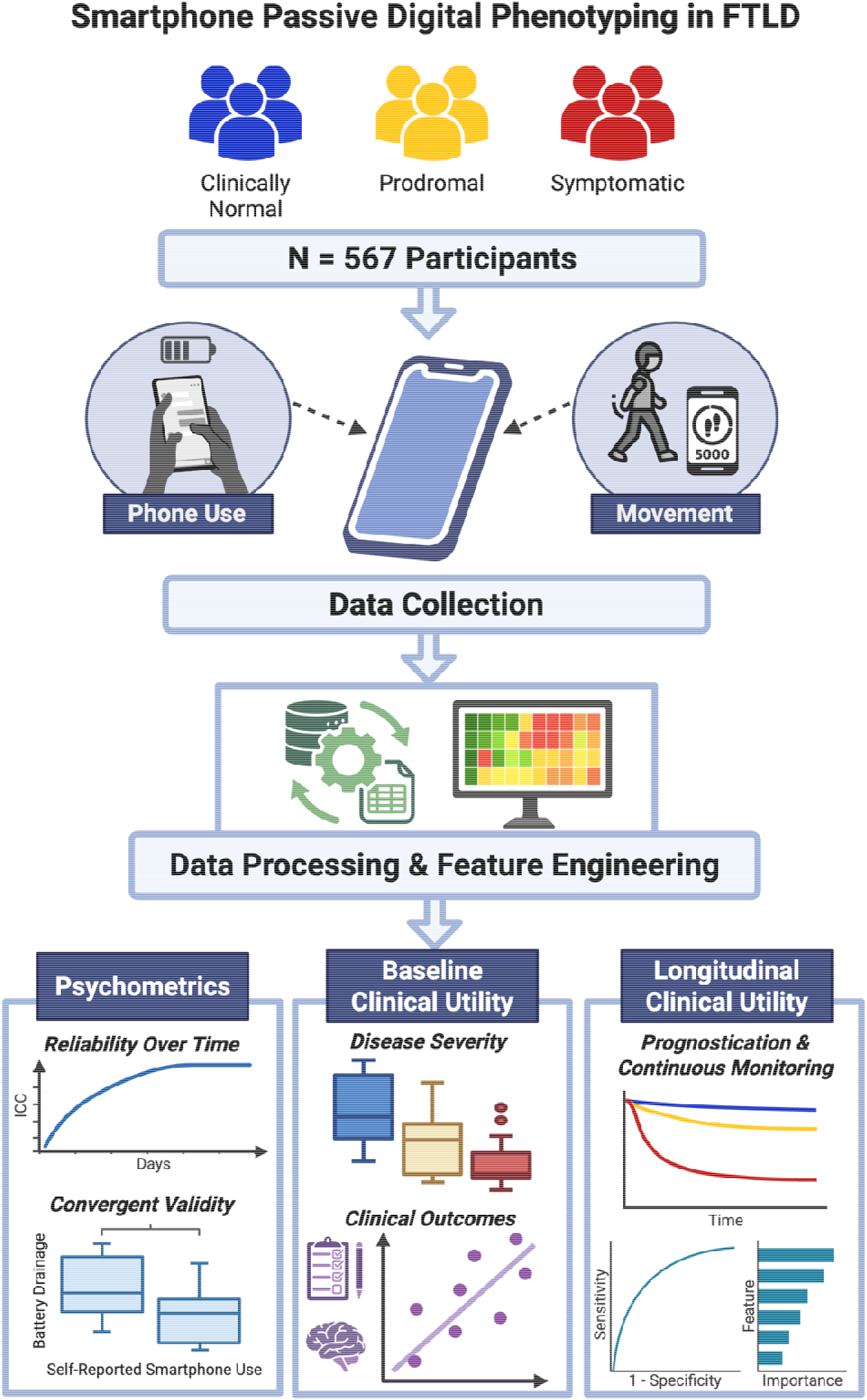
Study Overview. Data on smartphone use (battery drainage) and movement (step count) were collected through the ALLFTD Mobile App. Raw digital data were cleaned and processed, resulting in features that captured average daily and day-to-day variability in smartphone use and movement. This study evaluated psychometric properties and clinical utility of these passive smartphone features in frontotemporal lobar degeneration (FTLD).

## METHODS

### Participants

Participants included 567 adults recruited from parent studies of FTLD, including the ALLFTD Consortium (NCT04363684) and local studies of FTLD at UCSF (AG038791, AG062422, and AG019724). This study represents secondary analysis of data from the ALLFTD Mobile App Study, which has been described previously (20). Parent study participants included those who had a referring diagnosis of an FTLD clinical syndrome or who were members of a family with a strong family history of an FTLD syndrome. Additional inclusion criteria for this Mobile App Study were as follows: (1) aged ≥18 years, (2) access to a smartphone, and (3) English reported as the primary language. Participants were asked to use their own smartphones. Exclusion criteria were consistent with the parent ALLFTD study (20, 22).

Recruitment primarily targeted those with Clinical Dementia Rating plus National Alzheimer’s Coordinating Center FTLD module (CDR^®^+NACC-FTLD) Global scores of <2, but participants who were more severely impaired were not excluded. Data for this study were collected from August 2020 to August 2025. The study was approved by a centralized single institutional review board at Johns Hopkins Medicine (IRB # 20-29891), and all participants or legally authorized representatives provided written informed consent.

### Procedures

Participants were recruited to the ALLFTD Mobile App Study within 90 days of their in-person parent study visit during which they completed a comprehensive neurobehavioral evaluation, including clinical interview, neurologic examination, cognitive testing, neuroimaging, and study partner / informant interview. Once enrolled, participants downloaded the ALLFTD Mobile App (available on both iOS and Android devices) onto their personal smartphones with the assistance of a site clinical research coordinator. Participants then completed all cognitive tests on the app without supervision, as described previously (20). The app collected continuous passive data on battery percentage and step count as long as the app remained on participants’ phones. Longitudinal follow-up was completed via smartphone monitoring and annual in-person study visits.

### Measures

#### Passive Smartphone Data

Daily phone use was estimated via proxy assessment of daily battery drainage, which despite noted limitations, has been validated previously in relation to neurocognitive outcomes in FTLD (28). To derive daily battery drainage, hourly decreases in battery percentages were summed across each 24-hour day (i.e., 00:00 to 23:59 local time).

Days with greater battery drainage are assumed to reflect greater smartphone use. Daily step counts were sourced directly from each phone’s built-in step tracker. iOS devices provide total step counts in 15-minute intervals, whereas Android devices provide total step counts during intervals when activity is detected. Step counts were summed within each 24-hour day. Days with less than 10 hours of battery or step count data captured were excluded. At baseline, herein defined as the first 30 days of smartphone monitoring, daily averages were calculated by taking the mean of daily values. Day-to-day variability was estimated by calculating the root mean square of successive differences (RMSSD) from daily values, which importantly accounts for variability across consecutive days. Participants with fewer than 4 days of data represented at baseline were considered to have too much missing data for reliable quantification of baseline smartphone use and step count and were therefore excluded from baseline analyses. To capture longitudinal changes in these passive smartphone features over time, average and day-to-day variability (RMSSD) in smartphone use and step count were calculated within each week of their longitudinal study participation. Aggregate average and variability estimates for step count data were log-transformed to achieve approximate normality for analysis.

*Mobile Cognitive Testing.* Participants completed five self-administered gamified cognitive tests of memory and executive functioning on their own smartphones, which were used to derive a global cognitive composite score (25). These included: 1) an adaptive memory task (Humi’s Bistro; primary metric = mean correct across trials), 2) flanker (Ducks in a Pond; primary metric = combined speed-accuracy score), 3) Stroop (Color Clash; primary metric = combined speed-accuracy score), 4) 2-back (Animal Parade; primary metric = d-prime, a combination of hits and false positives), and 5) card sort (Card Shuffle; primary metric = total correct). Detailed scoring procedures for each of these tasks has been described elsewhere (20). Scores from each task were then z-scored and averaged together to calculate the App global composite score, requiring data from at least 2 tests to compute. Participants also completed a Technology Familiarity questionnaire through the App, which assessed self-reported smartphone use (daily vs. less than daily), confidence in using their smartphone (confident vs. less than confident), and phone proximity/carriage (binarized to those who reported “always” keeping phone with them vs. those who reported anything less than “always”).

#### Annual In-Person Cognitive

*Functional, Neurologic, and Neuropsychiatric Assessment.* Participants completed the Uniform Data Set (UDS) version 3 (33) battery of cognitive tests as part of their parent ALLFTD study visit. The UDS version 3 Executive Function (UDS3-EF) composite score was used as a primary measure of executive functioning, comprising scores from the Trail Making Test, phonemic fluency, category fluency, and number span backwards (34). The UDS3 Memory composite score (35) was used as a primary measure of memory, comprising scores from the Montreal Cognitive Assessment (MoCA) Delayed Recall, Benson Figure Delayed Recall, Craft Story Immediate and Delayed Recall, and California Verbal Learning Test version 3 Brief Form (CVLT-3-BF) Immediate Recall Trials 1-4 and Delayed Free Recall. Language functioning was assessed via the Multilingual Naming Test (MINT) (36). Visuospatial functioning was assessed via the Benson Complex Figure Copy (37). Informant and participant interviews were conducted to derive CDR^®^+NACC-FTLD Global and Sum of Boxes scores (38). CDR^®^+NACC-FTLD Global scores were used to categorize participants as clinically normal (0), prodromal (0.5), or symptomatic (≥1). Participants’ motor functioning was evaluated via the Progressive Supranuclear Palsy Rating Scale (PSPRS), completed by a neurologist during the neurologic examination (39). The subset of PSPRS items comprising the limb-motor subscale (possible score range = 0-16), gait-midline subscale (possible score range = 0-20), and history of falls (possible score range = 0-5) were used for this study to identify those with any motor symptoms (i.e., sum of included items ≥1 vs. 0). Informants also completed the Neuropsychiatric Inventory Questionnaire (NPI-Q) (40) to assess the presence and severity of neuropsychiatric symptoms in participants. NPI-Q total severity score was used for this study, quantified by summing the severity ratings from each of the 12 domains assessed (possible range = 0-36).

#### Neuroimaging

T1-weighted images from 3T MRI scanners were acquired as Magnetization Prepared Rapid Gradient Echo images using the following parameters: 240 × 256 × 256 matrix; about 170 slices; voxel size = 1.05 × 1.05 × 1.25 mm3; flip angle, echo time and repetition time varied by vendor. A standard imaging protocol was used across all centers and all images were reviewed for quality by a core group at the Mayo Clinic, Rochester. Details of image acquisition, processing, and harmonization have been published elsewhere (41). Briefly, SPM12 (Statistical Parametric Mapping) was used for segmentation (42), Large Deformation Diffeomorphic Metric Mapping was used to generate a customized group template (43), and regional volumes were extracted using the Desikan atlas (44). The primary volumetric neuroimaging variable of interest was total gray matter volume as a biomarker of neurodegeneration, which was residualized on total intracranial volume using simple linear regression to account for inter-individual differences in head size.

### Statistical Analysis

All analyses were completed using R, version 4. To characterize the sample by disease severity, differences in demographics, clinical characteristics, and passive smartphone features by CDR^®^+NACC-FTLD Global score groups (0 vs. 0.5 vs. 1+) were tested with one-way ANOVA or chi-square tests for continuous or categorical variables, respectively. Differences in demographics and smartphone passive features by device type (iOS vs. Android) were evaluated with independent t-tests. Reliability of passive smartphone features was evaluated by the intraclass correlation coefficient (ICC) per the Shrout and Fleiss 1979 method for evaluating test-retest reliability from the mean of measurements (45, 46), and calculated as var_B_/(var_B_ + var_W_/days), where var_B_ is the between subject variance, and var_W_, the within subject between days variance were estimated using a random effects model and days is the number of days of data used for calculation of the smartphone feature measure. ICC values are interpreted based on published suggestions (poor: < 0.5; moderate: 0.5-0.75, good: 0.75-0.9; excellent: >0.9) (45).

Convergent validity of smartphone use estimates by self-reported smartphone use was evaluated with independent t-tests. Pearson correlations examined associations between passive smartphone features and continuous measures of cognitive functioning, neuropsychiatric symptoms, and brain volumes. Linear regression also examined these associated while covarying for demographics (age, sex, and years of education) and device type (iOS vs. Android). Linear mixed-effect models (“lme4” (47) and “lmerTest” (48)) were used to test all longitudinal associations, including models examining baseline smartphone passive features as predictors of longitudinal trajectories of functional decline and models examining functional change (last minus first CDR^®^+NACC-FTLD Sum of Boxes score) as a predictor of longitudinal trajectories of passive smartphone features. All linear mixed-effects regressions modeled person-specific random intercepts and random effects of time (quantified as years since baseline).

Random forest classification was used to evaluate incremental validity of adding passive smartphone features to the app-based global cognitive composite score for prediction of functional decline. All models included baseline age, sex, and years of education. Per published recommendations for relatively small biomedical datasets, nested cross validation was utilized, i.e., 5-fold cross-validation in an inner loop for hyperparameter tuning and separate 5-fold cross-validation in an outer loop for model testing (49–52). Random forest analyses were completed using the “ranger” (53) and “nestedcv” packages (51). Model performances were evaluated by pooling the predicted probabilities from hold out samples of outer loop folds to calculate areas under the curve (AUC) utilizing the entire sample for each model. Model comparisons were then performed by comparing model AUCs via paired DeLong tests. Given that this model comparison method may introduce bias by combining predictions from distinct refits calibrated on different training sets, two additional sensitivity analyses were performed: 1) we evaluated AUCs for each of the hold out samples and compared models using paired t-tests, which removes the bias from pooling refits but has reduced efficiency; and 2) we fit classic logistic regression models on the entire sample and compare these nested models via likelihood ratio tests.

## RESULTS

### Participant Characteristics

In the full sample of 567 participants (**Table 1**), over half were female (57%), a majority self-identified as non-Hispanic White (95%), and average (SD) age was 52.8 (14.7) years. Participants had 16 years of education on average (SD = 2.4). Participants were followed across 2.6 annual in-person study visits on average (SD = 1.3; range = 1-5). Among the 554 participants with available CDR^®^+NACC-FTLD (38) scores at baseline, 295 (53%) were clinically normal (global score = 0), 117 (21%) were prodromal (global score = 0.5), and 142 (26%) had at least mild dementia (i.e., “symptomatic”; global score ≥ 1). Among the 443 (78%) participants with available genetic testing results, 186 carried a pathogenic familial FTLD variant, including *GRN* (n=37), *MAPT* (n=45), or the *C9orf72* repeat expansion (n=92).

**Table 1.** Demographic and Clinical Characteristics.

|  | Full Sample<br>(N = 567) | A.<br>Clinically<br>Normal<br>(N = 295) | B.<br>Prodromal<br>(N = 117) | C.<br>Symptomatic<br>(N = 142) | p | Pairwise |
| --- | --- | --- | --- | --- | --- | --- |
| *Demographics* |  |  |  |  |  |  |
| Age | 52.8 (14.7) | 45.0 (13.3) | 60.2 (11.6) | 63.7 (8.5) | <.001 | A<B<C |
| Sex (female) | 324 (57%) | 197 (67%) | 56 (48%) | 61 (43%) | <.001 | A>B,C |
| Education | 16.2 (2.4) | 16.3 (2.3) | 16.2 (2.5) | 15.9 (2.6) | .28 |  |
| Race (White) | 538 (95%) | 280 (95%) | 114 (97%) | 133 (94%) | .36 |  |
| *Study Characteristics* |  |  |  |  |  |  |
| Device Type |  |  |  |  | .71 |  |
| iOS | 406 (72%) | 206 (70%) | 86 (74%) | 103 (73%) |  |  |
| Android | 161 (28%) | 89 (30%) | 31 (26%) | 39 (27%) |  |  |
| Total in-person study visits | 2.6 (1.3)<br>[range = 1-5] | 2.8 (1.2) | 2.5 (1.3) | 2.2 (1.2) | <.001 | A>C |
| Days of passive data captured | 522.9 (448.0)<br>[range = 4-1859] | 582.7(472.8) | 536.8<br>(443.9) | 405.8<br>(377.3) | .002 | A>C |
| *Clinical Characteristics* |  |  |  |  |  |  |
| Genetic Status |  |  |  |  |  |  |
| Results Available | 443 (78%) | 249 (84%) | 89 (76%) | 102 (72%) | .006 | A>C |
| Carrier of Pathogenic Variant | 186 (33%) | 134 (45%) | 33 (28%) | 17 (12%) | <.001 | A>B>C |
| *C9orf72* | 92 (16%) | 65 (22%) | 19 (16%) | 6 (4%) |  |  |
| *MAPT* | 45 (8%) | 27 (9%) | 12 (10%) | 6 (4%) |  |  |
| *GRN* | 37 (7%) | 32 (11%) | 2 (2%) | 3 (2%) |  |  |
| Other<sup>a</sup> | 12 (2%) | 10 (3%) | 0 (0%) | 2 (1%) |  |  |
| Clinical Phenotype<sup>b</sup> |  |  |  |  | - |  |
| Clinically normal | 290 (51%) | 283 (96%) | 6 (5%) | 0 (0%) |  |  |
| MCI/MCBI | 47 (8%) | 0 (0%) | 46 (39%) | 0 (0%) |  |  |
| bvFTD | 88 (16%) | 0 (0%) | 18 (15%) | 70 (49%) |  |  |
| nfvPPA | 27 (5%) | 0 (0%) | 14 (12%) | 13 (9%) |  |  |
| svPPA | 36 (6%) | 0 (0%) | 8 (7%) | 28 (20%) |  |  |
| lvPPA | 1 (<1%) | 0 (0%) | 0 (0%) | 1 (1%) |  |  |
| PSPS | 20 (4%) | 0 (0%) | 6 (5%) | 14 (10%) |  |  |
| CBS | 13 (2%) | 0 (0%) | 7 (6%) | 6 (4%) |  |  |
| FTD/ALS or ALS | 10 (2%) | 0 (0%) | 3 (2%) | 6 (4%) |  |  |
| Other<sup>c</sup> | 23 (4%) | 9 (3%) | 10 (9%) | 4 (3%) |  |  |

Note. Full Sample includes all participants, including 13 who did not have an available CDR+NACC-FTLD rating. Clinically normal includes adults without functional impairment (CDR+NACC-FTLD Global = 0) who may be carriers of pathogenic variants. Prodromal is defined as CDR+NACC-FTLD Global = 0.5. Symptomatic participants include anyone with CDR+NACC-FTLD Global ≥ 1. MCI = mild cognitive impairment; MCBI = mild cognitive behavioral impairment; bvFTD = behavioral variant frontotemporal dementia; nfvPPA = nonfluent variant primary progressive aphasia; svPPA = semantic variant PPA; lvPPA = logopenic variant PPA; PSPS = primary progressive supranuclear palsy syndrome; CBS = corticobasal syndrome; ALS = amyotrophic lateral sclerosis.
<sup>a</sup>Includes identified rare pathogenic variants not specified to protect participant anonymity
<sup>b</sup>N=555 participants had available data for clinical phenotype at baseline
<sup>c</sup>Includes clinical phenotypes that don't meet criteria for syndromes listed in the table, including amnestic-predominant dementia syndromes, Parkinson's disease, late life psychiatric disorders, or unspecified.

Most participants used iOS devices (72%; versus 28% Android). See Supplementary Table 1 for all comparisons between iOS and Android users at baseline. In brief, iOS and Android users were comparable by baseline age, sex, and education (ps > 0.05; Suppl. Table 1). iOS users had lower estimates of average daily drainage in battery percentage (mean = 40% per day) compared to Android users (mean = 49% per day; p = 0.001; Suppl. Table 1). Estimated differences in average step counts, day-to-day variability in step counts, and day-to-day variability in battery drainage between iOS and Android users, however, did not reach statistical significance (ps > 0.05).

### Feasibility

Out of all 567 participants who enrolled and logged into the app, only 10 (2%) had too much missing passive data at baseline (defined as less than 4 days of data captured) to reliably quantify passive smartphone features of interest, including daily smartphone use or step counts. Rates of missing data did not meaningfully differ by disease severity (2% missingness in clinically normal; 2% missingness in prodromal participants; 2% in symptomatic participants; *p* = 0.95); however, all 10 participants with missing passive data were using iOS devices (versus Android, *p* = 0.10). Notably, all 63 (100%) participants who logged in but did not complete any of the five mobile cognitive tests at baseline had smartphone passive data available, representing an 11% increase in smartphone data availability.

### Reliability of Passive Smartphone Features

To evaluate the reliability of our estimates of average daily smartphone use and step count, we estimated the intraclass correlation coefficients (ICCs) as a function of the number of days of passive data used in the calculation of the smartphone feature, from 2 to 30 days, with the baseline period being at most 30 days. For smartphone use, only 3 days of battery data were needed to achieve good reliability (ICC = 0.77, 95%CI [0.73-0.80]), whereas excellent reliability was achieved with 8 days (ICC = 0.91, 95%CI [0.89-0.92]) of monitoring (**Fig. 2A**). For step counts, good reliability was achieved within 4 days (ICC = 0.76, 95%CI [0.72-0.79]) while excellent reliability was achieved within 11 days (ICC = 0.91, 95%CI [0.89-0.92]) of monitoring (**Fig. 2B**). For the clinical use case of capturing within-person change over longitudinal follow-up and to identify minimal detectable levels of change in the passive smartphone features, we also characterized person-specific standard errors of measurement based on data from the entire 30-day baseline period. For example, person-specific standard errors could be used as a threshold to flag potentially meaningful change in a passive smartphone feature beyond measurement error. Person-specific standard errors in battery drainage (i.e., our proxy of smartphone use) had a median of 3.7 [IQR = 2.7 to 5.0]. Person-specific standard errors in step count had a median of 391.8 [IQR = 247.1 to 575.0].

**Figure 2.**
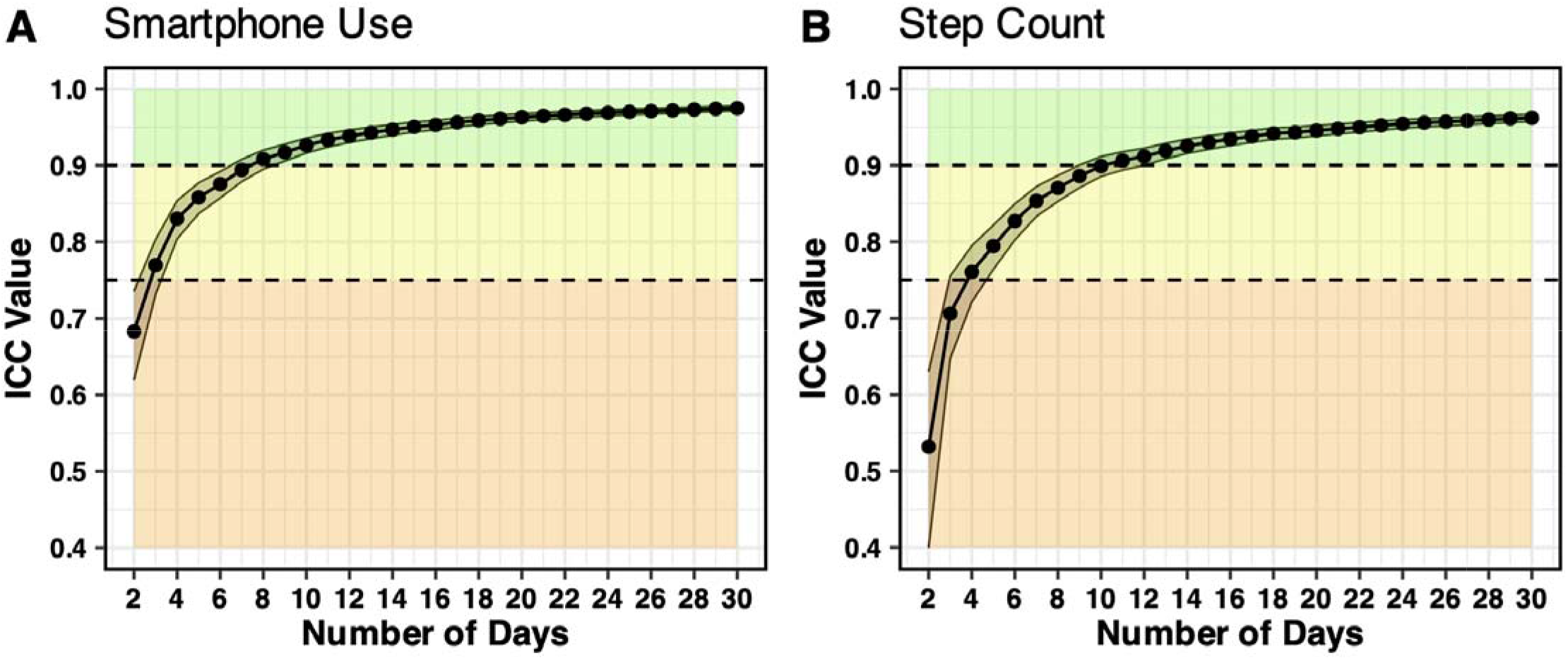
Reliability by Study Day. To evaluate reliability of (A) smartphone use and (B) step count features, intraclass correlation coefficients (ICCs) were estimated as a function of the number of days of smartphone monitoring data (using the mean of daily measurements). We demonstrate that only 3 days of smartphone use data and 4 days of step count data were needed to achieve good (ICC>0.75) reliability, while excellent (ICC>0.9) reliability was achieved with 8 and 11 days of smartphone use and step count data, respectively. Error bands represent 95% confidence intervals around ICC estimates.

### Agreement with Self-Reported Smartphone Use

To demonstrate validity of our estimate of average smartphone use (i.e., total battery drainage per day, on average), we examined associations with self-reported smartphone use at baseline. Participants who reported using their smartphone once per day or less (n=33) had statistically significantly lower estimates of daily smartphone use at baseline (mean = 27.3% battery used per day, SD = 17.6) compared to those who reported using their smartphone multiple times per day or more (mean = 45.2% battery used per day, SD = 21.5; mean difference = 17.9, 95%CI [10.8 to 24.5]; *p* < 0.001). Lower estimates of smartphone use were also observed among those who reported being “less than confident” in using their smartphone (mean = 36.1% battery used per day, SD = 22.1; versus “confident”: mean = 46.9% battery used per day, SD = 20.8; mean difference = 10.8, 95%CI [6.2 to 15.3]; *p* < 0.001) and those who reported keeping their phone on them less than “always” (mean = 36.5% battery used per day, SD = 19.8; versus “always”: mean = 45.4% battery used per day, SD = 19.7; mean difference = 8.9, 95%CI [1.4 to 16.3]; *p* = 0.021). All associations with self-reported smartphone use held in the subset of symptomatic participants.

### Passive Smartphone Features Associate with Clinical Factors at Baseline

CDR^®^+NACC-FTLD Global Score (0 vs. 0.5 vs. ≥1) was statistically significantly associated with all passive smartphone features at baseline (**Fig. 3A-3D**), such that those with more severe functional impairment had lower average smartphone use (48.0%/day vs. 41.2%/day vs. 33.2%/day; omnibus *p* < 0.001) and step counts (3985 steps/day vs. 3160.6 steps/day vs. 3147 steps/day; log-transformed for analysis, omnibus *p* < 0.001), as well as lower day-to-day variability (RMSSD) in smartphone use (25.0 vs. 21.5 vs. 21.4, omnibus *p* = 0.004) and step counts (3240.6 vs. 2535.4 vs. 2611.8; log-transformed for analysis, omnibus *p* < 0.001). Regarding cognitive functioning, baseline passive smartphone features demonstrated statistically significant correlations with both app-based and in-person traditional cognitive testing performance (**Fig. 3E**; see Supplementary Figure 1 for correlations among all variables represented). Although all passive smartphone features correlated with measures of executive functioning and memory, smartphone use features correlated more strongly with an in-person measure of language while step count features correlated more strongly with an in-person measure of visuospatial skills.

**Figure 3.**
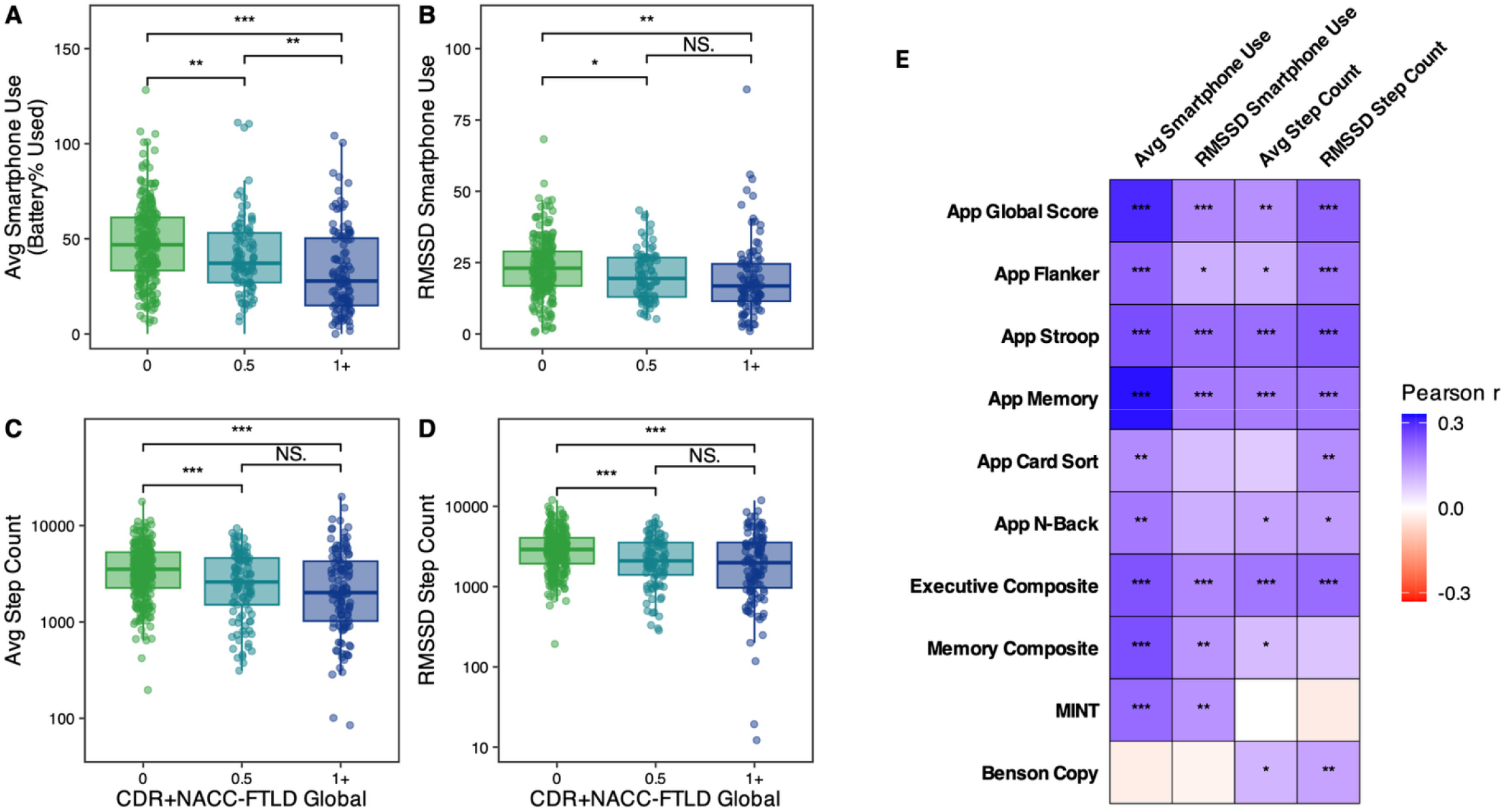
Baseline Associations with Functional Impairment and Cognitive Performance. Baseline functional impairment measured via the CDR+NACC-FTLD associated with baseline (A) average smartphone use, (B) day-to-day variability in smartphone use, (C) average step count, and (D) day-to-day variability in step count. (E) All four baseline passive smartphone features positively associated with global cognitive performance on the app, including measures of executive functioning and memory. While all four of these features also positively associated with in-person traditional measures of executive functioning, features characterizing average and variability in smartphone use were more strongly related to memory and language (Multilingual Naming Test [MINT]). In contrast, features characterizing average and variability in step count positively related to visuospatial skills (Benson Copy).

Participants with limb-motor symptoms, gait-midline symptoms, or a history of falls (yes/no) based on corresponding items from the clinician-rated PSPRS also had lower average smartphone use (*mean difference* = −8.0, 95%CI [−13.1 to −2.8], *p* = 0.001) and step counts (*mean difference* = −1394 steps, 95%CI [−1900 to −887], *p* < 0.001), as well as lower day-to-day variability in smartphone use (*mean difference* = −4.1, 95%CI [−6.7 to −1.4], *p* = 0.001) and step counts (*mean difference* = −980 steps, 95%CI [−1346 to −614], *p* < 0.001). All baseline passive smartphone features had small yet statistically significant correlations with neuropsychiatric symptoms (NPI-Q Total Score), such that more severe symptoms were associated with lower average daily smartphone use (Pearson *r* = −0.15, 95%CI [−0.25 to −0.06], *p* = 0.001), lower day-to-day variability in smartphone use (Pearson *r* = −0.10, 95%CI [−0.20 to −0.01], *p* = 0.040), lower average daily step count (Pearson *r* = −0.11, 95%CI [−0.20 to −0.02], *p* = 0.020), and lower day-to-day variability in step count (Pearson *r* = −0.15, 95%CI [−0.24 to −0.06], *p* = 0.001). In particular, participants with apathy had significantly lower average daily smartphone use (mean difference = −7.8, 95%CI [−12.4 to −3.2], p < 0.001), lower average daily step count (mean difference = −733.4, 95%CI [−1278.9 to −187.9], p = 0.009), lower day-to-day variability in step count (mean difference = −776.3, 95%CI [−1143.5 to −409.1], p < 0.001), and marginally lower day-to-day variability in smartphone use (mean difference = −2.3, 95%CI [−4.9 to 0.3], p = 0.084). Finally, among a subset of n=439 participants with volumetric neuroimaging data at their baseline visit, passive smartphone features correlated with total gray matter volumes (residualized on total intracranial volume), including average smartphone use (Pearson *r* = 0.26, 95%CI [0.16 to 0.35], *p* < 0.001), variability in smartphone use (Pearson *r* = 0.17, 95%CI [0.07 to 0.27], *p* = 0.001), average step count (Pearson *r* = 0.25, 95%CI [0.15 to 0.34], *p* < 0.001), and variability in step count (Pearson *r* = 0.26, 95%CI [0.16 to 0.35], *p* < 0.001). Associations held after covarying for demographics and device type (iOS vs. Android) in linear regression models.

### Passive Smartphone Features Prognosticate Rate of Functional Decline

In a linear mixed effect model covarying for baseline age, sex, education, and each of their interactions with time (years since baseline), baseline average smartphone use associated with trajectories of CDR^®^+NACC-FTLD Sum of Boxes (higher = more severe impairment; average smartphone use z-score x time in years: *b* = −0.201, *95%CI* [−0.35 to −0.06], *p* = 0.008; **Fig. 4A**). Baseline average smartphone use remained independently associated with functional trajectories even after adding the baseline app-based global cognitive score, baseline CDR^®^+NACC-FTLD Global score, and their interactions with time into the model (p < 0.05; **Table 2**). Similarly, baseline variability in smartphone use significantly associated with functional trajectories (RMSSD smartphone use z-score x time: *b* = −0.173, *95%CI* [−0.291 to −0.055], *p* = 0.004; **Fig. 4B**) and remained independent associated (p < 0.05) after adjusting for baseline app-based global cognitive score, baseline CDR^®^+NACC-FTLD Global score, and their interactions with time (see Supplementary Table 2). Baseline average daily step count (average step count z-score x time: b = −0.211, *95%CI* [−0.375 to −0.047], *p* = 0.012; **Fig. 4C**) and baseline day-to-day variability in step count (RMSSD step count z-score x time: b = −0.245, *95%CI* [−0.402 to −0.088], *p* = 0.002; **Fig. 4D**) were also significantly associated with functional trajectories in separate models; however, after adding baseline app-based global cognitive score, baseline CDR^®^+NACC-FTLD Global score, and their interactions with time, their regression estimates were notably reduced and did not reach statistical significance (see Supplementary Tables 3-4).

**Figure 4.**
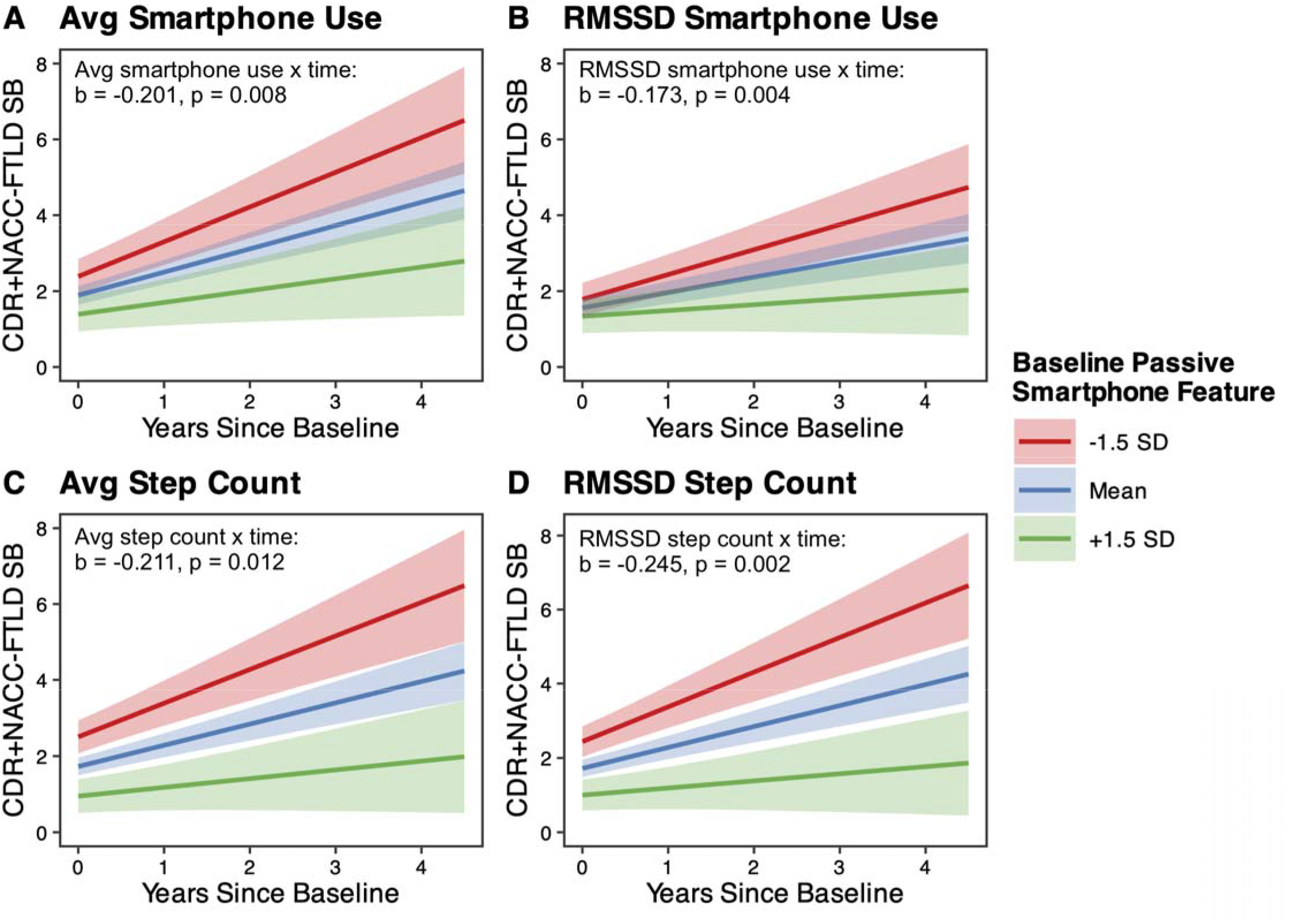
Baseline Passive Smartphone Features Prognosticate Decline. Distinct colors represent low. (red = 1.5 SD below the sample mean), medium (blue = sample mean), and high (green = 1.5 SD above the sample mean) values of the four different passive smartphone features at baseline. All four passive smartphone features capturing (A) average (avg) smartphone use, (B) day-to-day variability in smartphone use, (C) average step count, and (D) day-to-day variability in step count during the first 30 days of monitoring significantly predicted rates of change on the CDR+NACC-FTLD Sum of Boxes (SB), a gold-standard measure of functional impairment, over time. Plotted slopes and 95%CIs are derived from linear mixed effects regressions that modeled person-specific random intercepts and slopes of time. Covariates included baseline age, sex, education, and each of their interactions with time.

**Table 2.** Baseline Passively Collected Smartphone Use Prognosticates Functional Decline (CDR+NACC-FTLD Sum of Boxes)

|  | Step 1: WITHOUT additional clinical covariates |  | Step 2: WITH additional clinical covariates |  |
| --- | --- | --- | --- | --- |
|  | *Estimate [95%CI]* | *p* | *Estimate [95%CI]* | *p* |
| Time (years since baseline) | 0.58 [0.40, 0.76] | <.001 | 0.42 [0.29, 0.56] | <.001 |
| Baseline average smartphone use | -0.33 [-0.59, -0.07] | .012 | 0.04 [-0.06, 0.15] | .408 |
| Baseline age | 1.22 [0.96, 1.48] | <.001 | -0.03 [-0.16, 0.09] | .600 |
| Sex (ref: male) | 0.92 [0.42, 1.42] | <.001 | -0.01 [-0.22, 0.20] | .926 |
| Education (years) | -0.28 [-0.51, -0.04] | .022 | 0.03 [-0.06, 0.13] | .503 |
| **Time x Average smartphone use** | **-0.20 [-0.35, -0.05]** | **.008** | **-0.12 [-0.23, -0.01]** | **.028** |
| Time x Age | 0.35 [0.20, 0.50] | <.001 | -0.21 [-0.25, -0.08] | .002 |
| Time x Sex (ref: male) | 0.07 [-0.22, 0.36] | .634 | 0.05 [-0.17, 0.27] | .659 |
| Time x Education | -0.19 [-0.32, -0.05] | .008 | 0.03 [-0.08, 0.13] | .584 |
| Baseline app global cognitive score | -- |  | -0.19 [-0.33, -0.05] | .009 |
| Baseline CDR+NACC-FTLD Global | -- |  | 2.37 [2.24, 2.51] | <.001 |
| Time x App global score | -- |  | -0.60 [-0.76, -0.44] | <.001 |
| Time x CDR+NACC-FTLD Global | -- |  | 0.27 [0.12, 0.42] | .001 |

*Note.* Values represent unstandardized betas (95%CI) from each linear mixed effects model. All continuous variables in the model were standardized, except time and the outcome (i.e., CDR+NACC-FTLD Sum of Boxes). The outcome of interest, Time x Average Smartphone Use, is bolded and remains significantly significant at both steps of the model.

### Longitudinal Changes in Passive Smartphone Features Track with Functional Decline

To understand the utility of passive smartphone monitoring for tracking longitudinal clinical change over time, we tested whether longitudinal trajectories of passive smartphone features associated with changes in the CDR^®^+NACC-FTLD. Annual change in CDR^®^+NACC-FTLD Sum of Boxes was calculated by subtracting participants’ score at their last study visit from their score at their first study visit, then dividing by the number of years from first to last visit. All continuously collected passive smartphone data from the same timeframe were utilized and aggregated per week to achieve reliable estimates of smartphone use and step count over time. Values were standardized (z-scores) for comparability across models. Separate linear mixed effect models estimated trajectories in each passive smartphone feature as a function of change in CDR^®^+NACC-FTLD Sum of Boxes, time (years since baseline), and their interaction, covarying for baseline age, sex, education, and each of their interactions with time. Results showed that greater increases in CDR^®^+NACC-FTLD Sum of Boxes (i.e., steeper functional declines) were associated with steeper decreases in average smartphone use (b = −0.068, *95%CI* [−0.112 to −0.024], *p* = 0.003, marginal R^2^ = 0.16; **Fig. 5A**), variability in smartphone use (b = −0.081, *95%CI* [−0.130 to −0.033], *p* = 0.001, marginal R^2^ = 0.10; **Fig. 5B**), average step counts (b = −0.057, *95%CI* [−0.095 to −0.019], *p* = 0.003, marginal R^2^ = 0.13; **Fig. 5C**), and variability in step counts (b = −0.051, *95%CI* [−0.090 to −0.012], *p* = 0.012, marginal R^2^ = 0.11; **Fig. 5D**).

**Figure 5.**
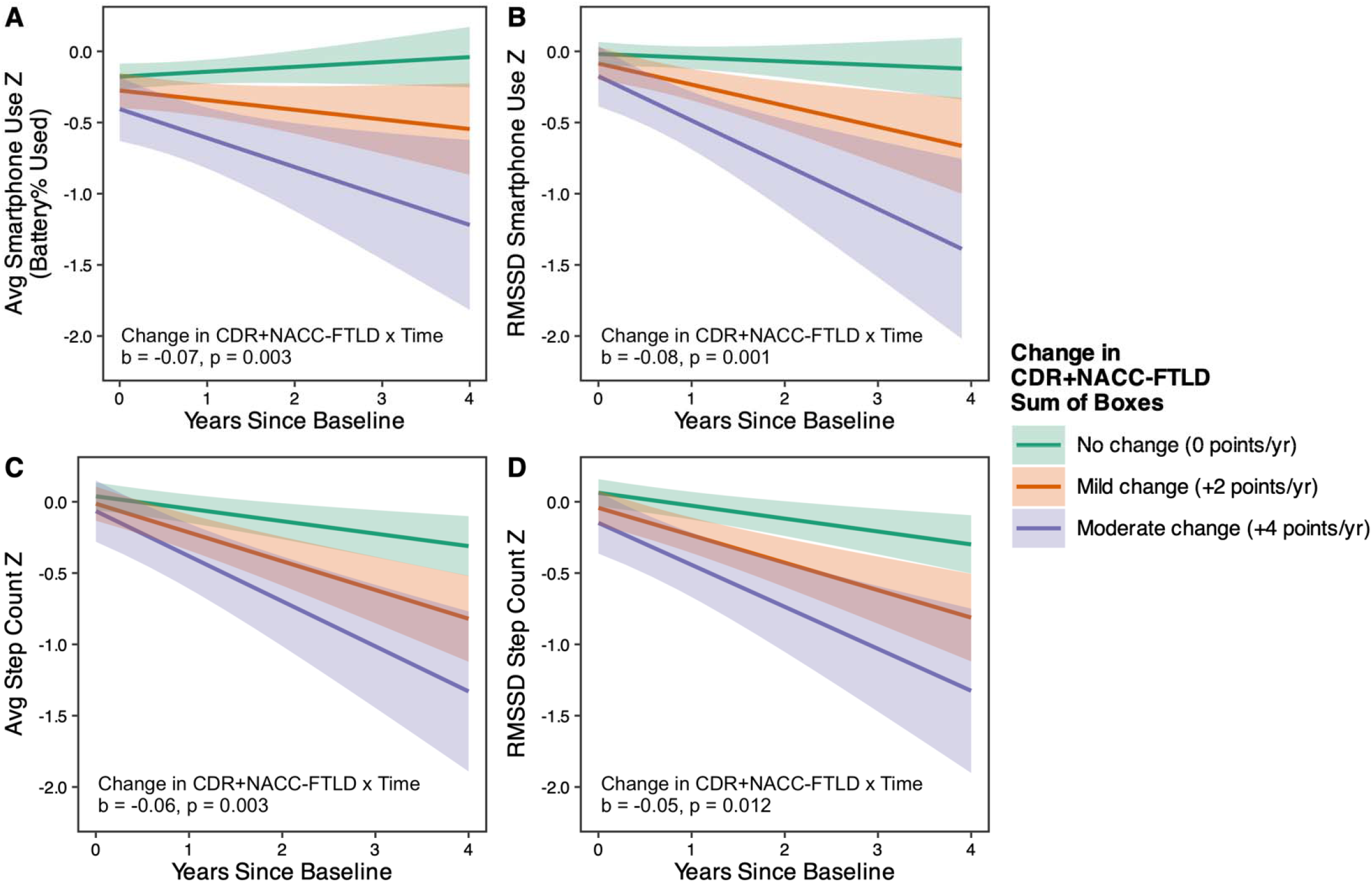
Real-Time Changes in Passive Smartphone Features Track with Functional Decline. Steeper functional declines from first to last study visit, as quantified by annual change in CDR+NACC-FTLD Sum of Boxes, associated with steeper declines in (A) weekly average (avg) smartphone use, (B) weekly variability in smartphone use, (C) weekly average step count, and (D) weekly variability in step count. Plotted slopes and 95%CIs are derived from linear mixed effects regressions that modeled person-specific random intercepts and slopes of time. Covariates included baseline age, sex, education, and each of their interactions with time.

### Passive Smartphone Features May Improve Detection of Functional Decline

We evaluated incremental validity of adding the passive smartphone features to a predictive model with the app-based global cognitive composite score to identify participants with evidence of clinical progression. The subset of 239 participants included in this analysis had at least 2 in person visits with CDR^®^+NACC-FTLD data to quantify functional changes, passive smartphone data over the same longitudinal timeframe, baseline app-based cognitive testing, and a baseline in-person traditional cognitive screener (MoCA). Controls (n=146, 61%) were defined as participants who were clinically normal at baseline *and* had a stable CDR^®^+NACC-FTLD Sum of Boxes score of 0 over time. Cases (n=93, 39%) were defined as participants who were impaired at baseline *or* had an increase in CDR^®^+NACC-FTLD Sum of Boxes; this also ensured capture of participants who converted from clinically normal to prodromal or symptomatic across the follow-up period. To anchor this analysis, we also evaluated the predictive performance of baseline MoCA score to classify these groups. Using 5-fold nested cross-validation, we trained and evaluated four random forest classification models: Model 1) MoCA plus demographics (age, sex, education); Model 2) App global score plus demographics; Model 3) Passive smartphone features plus demographics; Model 4) App global score and passive smartphone features plus demographics. The six passive smartphone features included in Models 3 and 4 were average and variability in smartphone use and step count across the entire monitoring period, as well as changes in average smartphone use and step count over time. AUCs for each model were calculated from pooled hold-out samples.

As seen in **Fig. 6A**, Model 1 (MoCA plus demographics) had an AUC of 0.85; Model 2 (App global score plus demographics) had an AUC of 0.86; Model 3 (passive smartphone features plus demographics) had an AUC of 0.84; and the combined Model 4 with App global score, passive smartphone features, and demographics had an AUC of 0.89. Feature importance and directionality of features in Models 3 and 4 are shown in **Fig 6B and Fig. 6C**, respectively, which highlight the app global cognitive composite score as an important feature in the final model and demonstrate the overall consistency of passive smartphone feature importance across models. Paired DeLong tests comparing AUCs from pooled hold-out sample predictions across models showed that Model 4’s AUC of 0.89 was statistically significantly higher than all other models (versus Model 1: AUC difference = 0.04, 95%CI [0.004 to 0.077], *p* = 0.029; versus Model 2: AUC difference = 0.03, 95%CI [0.002 to 0.055], *p* = 0.036; versus Model 3: AUC difference = 0.05, 95%CI [0.018 to 0.076], *p* = 0.001). Notably, Model 3 (passive smartphone features plus demographics) performance was comparable to that of the MoCA (Model 1; AUC difference = −0.01, 95%CI [−0.042 to 0.028], *p* = 0.709) and App global score (Model 2; AUC difference = −0.02, 95%CI [−0.056 to 0.018], *p* = 0.321) models. Given the lack of validated methods for nested cross validated model comparison, sensitivity analyses were performed to test other methods of classification and model comparison between the full and reduced models (see *Statistical Analysis*). First, paired t-tests comparing model performance (AUCs) for each hold-out sample across models showed that Model 4’s classification performance was statistically significantly better than Model 1 (*p* = 0.01) and Model 3 (*p* = 0.01) but not Model 2 (*p* = 0.42). Similarly, nested logistic regression models (i.e., identical predictors as Models 1-4) performed on the entire sample and compared via likelihood ratio tests showed that Model 4 was statistically significantly better than Model 1 (*p* = 0.04) and Model 3 (*p* < 0.01) but not Model 2 (*p* = 0.31).

**Figure 6.**
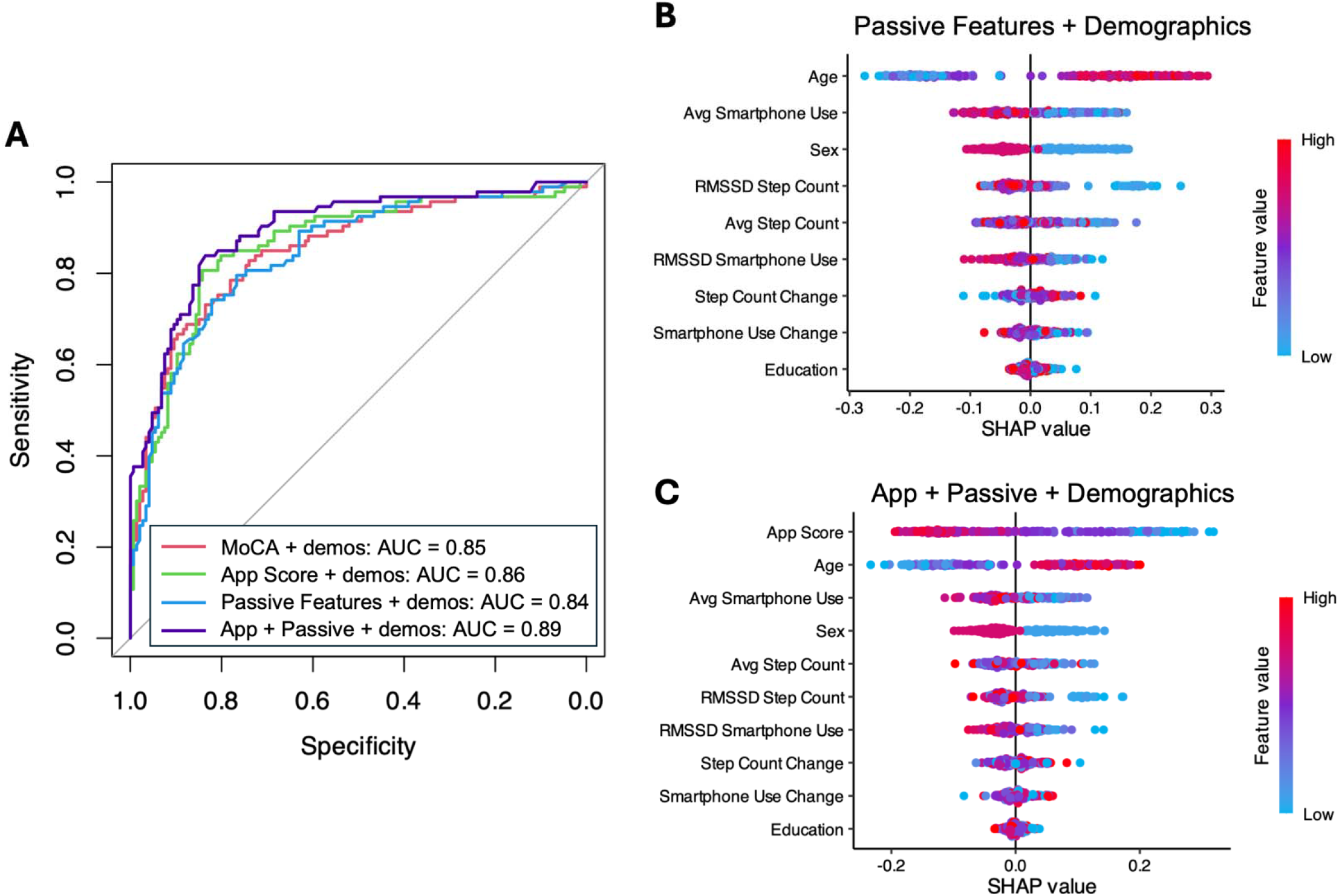
Passive Smartphone Features Improve Prediction of Longitudinal Functional Decline. (A) Receiver operating characteristic (ROC) curves are based on random forest classification models using 5-fold nested cross validation to identify participants who had functional decline over time vs. stably normal controls. The full model (purple) included the smartphone app global cognitive score, all passive smartphone features, and demographics (baseline age, sex, education) and had the highest AUC compared to all three other models (DeLong test *p*s < 0.05). (B, C) Beeswarm plots elucidate feature importance and directionality. Shapley additive explanations (SHAP) importance identified which features were most important for model predictions in Models 3 (passive features + demographics) and 4 (full model), ordered on the y-axes from most (top) to least (bottom) important. The color gradient shows directionality of each feature’s values (low = blue; high = red) in relation to the likelihood of functional decline (higher SHAP value = higher likelihood of functional decline), e.g., older age (represented in red) associated with greater likelihood of functional decline. Average smartphone use was the most important passive smartphone feature across both Models 3 and 4.

## DISCUSSION

We demonstrate that passive smartphone monitoring across the clinical spectrum of FTLD is feasible, reliable, valid, and clinically informative. Passive smartphone data were available for 98% of enrolled participants, even for those who had missing app based cognitive testing data. This high level of feasibility for passive data collection across stages of disease severity supports its potential for monitoring meaningful functional outcomes in persons whose illness is otherwise too progressed for engagement with objective cognitive testing. Further, passive smartphone data had excellent reliability, were associated with cognitive and functional outcomes at study baseline and longitudinally, and may add incrementally important information for functional monitoring beyond active cognitive testing, highlighting the value of passive monitoring both as an independent measure and as a complement to remote mobile cognitive testing. Together, these findings support applications of smartphone sensing technology to obtain low burden, unobtrusive monitoring of naturalistic behavior via smartphone sensing technology as a clinically valuable tool in FTLD.

These data strongly support the feasibility and reliability of passive smartphone data collected via the ALLFTD Mobile App. Only 3 days of smartphone use data and 4 days of step count data were needed to achieve good reliability, and 98% of participants met this threshold for data completeness. Given that these data are collected unobtrusively with no added burden to participants, passive smartphone monitoring represents a promising complementary mode of remote clinical measurement alongside mobile cognitive testing. This may be particularly useful for cases in which FTLD-related symptoms make it difficult to engage with cognitive testing. For example, the reliance on language for delivery of task instructions is a barrier for patients with more advanced primary progressive aphasia (PPA) syndromes even when the task itself assesses non-verbal functions (54, 55). Similarly, behavioral symptoms associated with behavioral variant FTD (bvFTD) such as apathy and disinhibition can also make it challenging to validly complete standardized cognitive tasks, especially when self-administered (20, 56). The completeness of passive smartphone data collected even among the participants with more severe impairment in this study support their feasibility across levels of disease severity.

The observed associations between passive smartphone features and gold-standard clinical measures of cognitive and functional impairment at baseline were consistent with our prior work (28) and with the hypothesis that those with more advanced FTLD disease severity would have reduced engagement with technology and movement (57–60). Although this monotonic association was expected with average smartphone use and step count, there have been varying hypotheses about how day-to-day variability in these behaviors changes with neurodegenerative disease progression. For example, some have proposed that day-to-day variability may increase in early stages of cognitive decline as patients recruit compensatory strategies to complete functional tasks less efficiently and more inconsistently, and then decrease again during the transition from mild cognitive impairment to dementia as overall levels of functioning decrease more drastically (61). Although we did not observe this non-linear pattern at the group level in our current data, we acknowledge individual cases in which identifying such non-linear patterns in any of the passive smartphone features may be clinically relevant, particularly in FTLD. Indeed, repetitive behaviors are one of the six diagnostic criteria of bvFTD, which commonly include specific behaviors such as pacing and repetitive phone use (62) that may be directly captured by passive smartphone monitoring. Future work is needed to explore syndrome specific patterns and develop methods that identify clinically meaningful change in behavior compared to one’s own baseline in real time, regardless of the direction of change, which could flag the need for clinical follow up.

Interestingly, passive smartphone features capturing smartphone use behavior appeared to be the strongest of the passive predictors of multi-domain cognitive and functional impairment across models. This may reflect the hypothesis that smartphone use, and technology use more broadly, is a cognitively complex task, making it a sensitive indicator of even subtle neurocognitive change (28, 57, 58). This hypothesis is further corroborated by previous studies showing that difficulty with everyday technology use is a common early functional concern in older adults with cognitive impairment that worsens with symptom progression (63, 64).

Similarly, while all the baseline passive features prognosticated future functional decline in models without other clinical covariates, only features capturing smartphone use behavior remained statistically significant predictors of functional trajectories independent of baseline mobile cognitive testing performance and disease severity (i.e., baseline CDR^®^+NACC-FTLD global score). These results support that quantifying use of personal digital devices may capture facets of everyday functional ability such as communication, planning, and task initiation, that are incompletely captured by cognitive testing data alone.

One of the major advantages of passive digital phenotyping is the continuous nature of data collection, allowing potential capture of neurocognitive changes as they are occurring in real time. Our findings support this potential, as we observed strong associations between longitudinal changes in passive smartphone features and rate of functional changes as assessed by gold standards over the same time frame. Notably, in predictive models, longitudinal passive smartphone features performed comparably to the MoCA and the app-based global cognitive composite score for detecting functional decline over study follow-up.

Further, including these passive smartphone features into a combined prediction model with the app-based global cognitive composite score and demographics improved classification performance; however, the size of this improvement in model performance was modest and not statistically significant in sensitivity analyses. Still, these continuous longitudinal passive digital data are supported as incrementally useful for monitoring FTLD disease progression independently and in addition to active smartphone-based cognitive assessments. Future work will examine important within-day phone use and activity patterns, e.g. (65), and incorporate other valuable data collected from the multi-domain assessments on the ALLFTD Mobile App, including from tasks that evaluate speech and motor functioning.

This study has several strengths, including being among the first to implement smartphone passive digital phenotyping in a large sample of adults with neurodegenerative disease; however, there are also some limitations. First, we demonstrated some variation in our proxy measure of smartphone use by device type with Android devices having greater battery drainage than iOS devices. Although analyses adjusted for device type and associations persisted, platform level differences and OS updates may influence absolute values. Further, while our use of battery drainage as a proxy for smartphone use allowed greater inclusion of iOS devices as opposed to more protected and inaccessible data such as screen time (28), we also acknowledge that battery drainage can be affected by non behavioral factors such as background processes, specific app use (e.g., maps/navigation), system settings, and battery health. Similarly, while we did not find statistically significant differences in step count feature by device type, there are known differences in step-count algorithms by platform. Future work is needed to characterize hardware- and software-specific differences and determine whether harmonization efforts are needed. Next, our cohort was predominantly non Hispanic White and primarily iOS users, which may limit generalizability. Studies are underway to extend this work to more representative samples. Further, disease-specificity cannot be determined from these data, as differential relationships by pathology-specific subgroups were not tested and non-FTLD neurodegenerative diseases (e.g., AD, Lewy body disease) were not included.

In summary, passive smartphone monitoring in FTLD is feasible, reliable even over short monitoring windows, valid, and sensitive to clinically meaningful indicators of disease progression. These data support efforts to incorporate passive smartphone monitoring alongside active remote assessments in longitudinal FTLD research to inform real-time changes in symptom manifestation and progression. With further validation and replication, these methods could also be implemented in clinical trials and clinical settings. Such clinical implementation would require careful attention to data governance and patient safety, including clear escalation protocols when change is identified, possible integration with existing clinical workflows to limit additional clinical burden, and transparency for patients and care partners about what is being collected and why. Given our robust validation results and support for clinical utility without any added patient burden, such careful and rigorous efforts towards broader implementation of passive monitoring are worthwhile.

## Data Availability

Qualified researchers can request data through the ALLFTD consortium website (https://www.allftd.org/data). In accordance with ALLFTD data sharing policies (https://www.allftd.org/policies), data will be made available to qualified researchers whose proposed use of the data has been approved.

## ACKNOWLEDGEMENTS

We acknowledge the invaluable contributions of the study participants and families as well as the assistance of the support staff at each of the participating sites.

## SOURCES OF FUNDING

Data collection and dissemination of the data presented in this paper were supported by the ALLFTD Consortium (U19 AG063911 (A.L.B., H.J.R. and B.F.B.), funded by the National Institute on Aging and the National Institute of Neurological Diseases and Stroke); the former ARTFL and LEFFTDS Consortia (ARTFL: U54 NS092089 (A.L.B., H.J.R. and B.F.B.), funded by the National Institute of Neurological Diseases and Stroke and National Center for Advancing Translational Sciences; LEFFTDS: U01 AG045390 (H.J.R. and B.F.B.), funded by the National Institute on Aging and the National Institute of Neurological Diseases and Stroke); the National Institute on Aging (NIA) grant R01 AG077557 (A.M.S.); the Association for Frontotemporal Degeneration; the Bluefield Project to Cure FTD; the Rainwater Charitable Foundation; and the Tau Consortium. The paper was reviewed by the ALLFTD Executive Committee for scientific content. Additional funding support to the UCSF MAC was provided by the National Institutes of Health (NIH) National Institute on Aging (NIA) grants P01 AG019724 (M.L.G.T, H.J.R., and W.W.S.), P30 AG062422 (G.D.R.), and R01 AG038791 (A.L.B.). This work is additionally supported by NIH grants K23 AG084883 (E.W.P.), P30 AG062677 (B.F.B.), R00 AG073453 (E.B.), U01 NS100620 (K.K. and B.F.B.), R01 AG058233 (S.L.); the Intramural Research Program of the NIH (program #: ZIAAG000935, ZIANS003154); the John F. Douglas Alzheimer’s Foundation (J.C.R); the Michael J. Fox Foundation; the Department of Defense; the Weston Brain Foundation; Brain Canada; and Workplace Safety Insurance Board.

The contributions of the NIH authors (J.Y.K., B.J.T., S.W.S.) were made as part of their official duties as NIH federal employees, are in compliance with agency policy requirements, and are considered works of the United States Government. However, the findings and conclusions presented in this paper are those of the authors and do not necessarily reflect the views of the NIH or the U.S. Department of Health and Human Services.

## Notes

### Competing Interest Statement

Authors EWP, SD, JCT, MSC, RF, RS, KBC, JHK, BLM, WWS, MLGT, SL, VS, EB, DC, CMC, RRD, ADV, ME, JAF, EDH, JYK, IRM, JCM, AP, BP, TP, KR, NR, AS, BW, WKK, LKF, and HWH declare no financial or non-financial competing interests.
BA receives research support from the Centers for Disease Control and Prevention, the National Institutes of Health (NIH), Ionis, Alector, Prion Alliance, and the CJD Foundation. He has provided consultation to Acadia, Ionis, Gate Biosciences, Regeneron, Nobelpharma, and Sangamo, and receives royalties from UpToDate. NG has participated or is currently participating in clinical trials of anti-dementia drugs sponsored by Bristol Myers Squibb, Eli Lilly/Avid Radiopharmaceuticals, Janssen Immunotherapy, Novartis, Pfizer, Wyeth, SNIFF (The Study of Nasal Insulin to Fight Forgetfulness) and the A4 (The Anti-Amyloid Treatment in Asymptomatic Alzheimers Disease) trial. She receives research support from Tau Consortium and the Association for Frontotemporal Dementia and is funded by the NIH. She consults for BCBSA. KK consults for Biogen, Eisai, and BioArctic with no personal compensation, received research support from Avid Radiopharmaceuticals and Eli Lilly, and receives funding from NIH and Alzheimers Drug Discovery Foundation. PAL has served as a site primary investigator for clinical trials sponsored by Alector, AbbVie, BMS, Transposon and Woolsey. His he receives research support from Biohaven. He receives research and salary support from the NIH-NIA and the Alzheimers Association-Part the Cloud partnership. CUO receives research funding from the NIH, Department of Defense, Association for Frontotemporal Degeneration, Lawton Health Research Institute, National Ataxia Foundation, Alector and Denali. He is also supported by the Robert and Nancy Hall Brain Research Fund, the Jane Tanger Black Fund for Young-Onset Dementias, and gifts from the Joseph Trovato Estate and the Minerva Foundation. He is also a consultant to AviadoBio, Syndeo, Alector Inc., Otsuka Pharmaceutical, Reata Pharmaceuticals, and Neuvivo. JCR declares consulting fees from Genentech and is a site PI for clinical trials sponsored by Eisai and Roche. GSD reports no competing interests directly relevant to this work. His research is supported by NIH (R01AG089380, U01AG057195, U01NS120901, U19AG032438, P30AG062677). He serves as a Topic Editor (Dementia) for DynaMed (EBSCO). He is a co-Project PI for a clinical trial in anti-NMDAR encephalitis, which receives support from NIH/NINDS (U01NS120901) and Amgen Pharmaceuticals. He has developed educational materials for Continuing Education Inc, and MJH Life Sciences. He owns stock in ANI Pharmaceuticals. His institution has received in-kind contributions for radiotracer precursors for tau-PET neuroimaging in studies of memory and aging (via Avid Radiopharmaceuticals, a wholly owned subsidiary of Eli Lilly). DJI receives research funding paid to the institution from the NIH, the Michael J. Fox Foundation, and the Lewy Body Dementia Association (LBDA); receives research funding paid to the institution for clinical trials by Alector, Cervo Med, Denali, Novartis, Passage Bio, and Prevail; and has unpaid positions on the scientific advisory board for the LBDA, the medical advisory board for the Association for Frontotemporal Degeneration, and the board of directors for the International Society for Frontotemporal Dementias. MCT has served as an investigator for clinical trials sponsored by Aribio, Roche/Genentech, Merck, UCB, Novo Nordisk and Janssen. She receives research support from the Brain Canada, Canadian Institutes of Health Research, National Insitute of Aging, Tanenbaum Institute of Science in Sport, Weston Brain Foundation, Workplace Safety and Insurance Board. She has consulted for Eisai, Lilly, Roche, UCB, Novartis and Novo Nordisk. SWS received research support from Cerevel Therapeutics (now part of AbbVie Inc.), and serves on the Scientific Advisory Committee of the Lewy Body Dementia Association, Mission MSA, and the GBA1 Canada Initiative. She also has patents pending on the use of plasma proteomics in the diagnosis of ALS and Parkinsons disease. BJT holds patents on the diagnostic testing and therapeutic implications of the hexanucleotide repeat expansion of C9orf72. He also holds patents and has patents pending on the use of plasma proteomics in the diagnosis of ALS and Parkinsons disease. BJT received research support from Cerevel Therapeutics (no part of AbbVie Inc.). JK has provided expert witness testimony in multiple pharmaceutical cases, acting for Janssen Biotech Inc, Apotex, HEC, Ezra, Puma Biotechnology, West-Ward Pharmaceuticals, and Teva Pharmaceuticals. He has also provided biostatistical consulting for Aviadobio Inc. BFB has served as an investigator for clinical trials sponsored by Alector, Cervomed, Cognition Therapeutics, and Transposon. He provides consulting services (no personal fees) to Acadia and GSK. He serves on the Scientific Advisory Board of the Tau Consortium which is funded by the Rainwater Charitable Foundation. He receives research support from NIH. HJR has received consulting fees from Eli Lilly, and receives research support from the NIH and the state of California. ALB is the lead principal investigator for both the NIH Alzheimers Disease Tau Platform (ATP) and the PSP Trial Platform (PTP), two new, multidrug public-private clinical trial partnerships designed to accelerate the development of effective therapies. He has initiated and led multiple public-private partnerships focused on accelerating drug development for neurodegenerative disease including the Neurofilament Surveillance Project, the FTD Treatment Study Group (now called the FTD Research Roundtable) and the PSP Research Roundtable. AMS received research support from the NIA/NIH, the Bluefield Project to Cure FTD, the ALS Association, the Association for Frontotemporal Degeneration. He has provided consultation to Alector, Aviado Bio, Cervomed, Coya, Otsuka, Prevail Therapeutics/Eli Lilly, Passage Bio, Takeda, and Vesper Bio.

### Author Declarations

A centralized single institutional review board at Johns Hopkins Medicine gave ethical approval for this work (IRB # 20-29891), and all participants or legally authorized representatives provided written informed consent.

